# Factors influencing the outcomes of non-pharmacological interventions for managing fatigue across the lifespan of people living with musculoskeletal (MSK) conditions: a scoping review protocol

**DOI:** 10.1101/2023.11.15.23298534

**Authors:** Katie Fishpool, George Young, Coziana Ciurtin, Fiona Cramp, Emmanuel Erhieyovwe, Bayram Farisogullari, Gary Macfarlane, Pedro M. Machado, Jen Pearson, Eduardo Santos, Emma Dures

## Abstract

**Introduction:** Fatigue is an important and distressing symptom for many people living with chronic musculoskeletal (MSK) conditions. Many non-pharmacological interventions have been investigated in recent years and some have been demonstrated to be effective in reducing fatigue and fatigue impact, however there is limited guidance for clinicians to follow regarding the most appropriate management options. The objective of this scoping review is to understand and map the extent of evidence in relation to the impact of non-pharmacological interventions on MSK condition-related fatigue across the lifespan.

**Methods and analysis:** This scoping review will include evidence relating to people of all ages living with chronic MSK conditions who have been offered a non-pharmacological intervention with either the intention or effect of reducing fatigue and its impact. Databases including AMED, PsycINFO, CINAHLPlus, MEDLINE, EMBASE and Scopus will be searched for peer reviewed primary research studies published after 1^st^ January 2007 in English language. These findings will be used to identify factors associated with successful interventions and to map gaps in knowledge.

**Ethics and dissemination:** Ethical approval was not required for this review. Findings will be disseminated by journal publication, conference presentation and by communicating with relevant healthcare and charity organisations.

**Article Summary:** *Strengths and limitations of this study:*

- Patient and Public Involvement and Engagement (PPIE) workshops at key time points will ensure that the protocol, review findings and subsequent discussion are relevant to stakeholders and reflect lived experience of MSK-fatigue
- All studies will be reviewed, and data extraction checked by a minimum of two researchers
- The effectiveness of specific interventions and methodological quality of included studies is not covered in this scoping review
- Only evidence available in English will be reviewed

## INTRODUCTION

Musculoskeletal (MSK) conditions include inflammatory and non-inflammatory conditions such as connective tissue diseases, inflammatory arthritis and osteoarthritis, back and neck pain, and fibromyalgia which affect the muscles, bones, joints and connective tissue (1,2). More than 10 million people in the UK and 1.7 billion people globally currently live with an MSK condition (1,2). Prevalence increases with age but these conditions are encountered across the lifespan, with approximately 234,000 children in England and Scotland living with a long term MSK condition (2).

Fatigue has been identified by people living with chronic MSK conditions as a priority symptom which has a significant impact on quality of life (2–8). Pharmacological treatments are not licensed for the management of fatigue without concurrent disease activity, so the focus in clinical practice has been on non-pharmacological options (9,10). This has been mirrored in healthcare research and recent systematic reviews have examined the strength of the evidence supporting a range of non-pharmacological interventions in different patient groups (11–15). Non-pharmacological interventions are any non-chemical or biological interventions that are theoretically based and empirically proven, or that have a logical rationale which is possible to prove by empirical study (11,16).

The European Alliance of Associations for Rheumatology (EULAR) recently funded a taskforce to examine the effectiveness of non-pharmacological interventions for fatigue in inflammatory rheumatic conditions and the resulting systematic review found strong evidence that some interventions are effective (14). This informed the EULAR recommendations for the management of fatigue (17) and highlighted a need to better understand contextual factors and the mechanisms by which interventions are effective (7). There is currently no comprehensive understanding of the factors which influence the success of an intervention. The impact of this is a lack of evidence to support decision making in how to design, offer and deliver interventions in the most effective way, tailored to a range of patients and at the optimal time. Current clinical guidelines for the management of common MSK conditions recognise fatigue as an important symptom but do not make any recommendations for how it can be addressed directly (18–20), hindering implementation of the evidence.

The clinical pathway for management of MSK conditions in the UK differs depending on primary diagnosis, with suspected inflammatory conditions being referred to specialist secondary care settings and osteoarthritis and fibromyalgia being managed predominantly through primary care (20–22). The impact of this is that the experience of patients with MSK fatigue and the profession and skills of the clinicians who provide their care may be quite different.

### Review aims

The aim of this scoping review is to generate evidence for health professionals and educators to design or adapt tailored MSK-fatigue support. The objectives of this review are to identify evidence for existing interventions for MSK-fatigue across the life course and to explore the theoretical basis for the interventions. To explore the comprehensive nature of the existing evidence, the clinical and demographic characteristics of the participants as well as to capture the training/skills of those who deliver the interventions where this information is available. As the intention is to create an overview of the current knowledge and to highlight gaps in the existing literature rather than to assess the effectiveness of specific interventions, a scoping review is the most appropriate methodological approach (23,24).

## METHODS AND ANALYSIS

### Study design

Recent and ongoing studies on the topic of fatigue support the need for this scoping review and have been used to inform its design. A previous scoping review exploring fatigue in patients with rheumatic and MSK conditions (25) reported on the efficacy of interventions and also considered determinants associated with fatigue. A systematic review assessed the quality of evidence available to support non-pharmacological interventions to reduce fatigue in patients with inflammatory rheumatic conditions (14). The National Institute for Health Research (NIHR) have recently awarded funding for the project “Effectiveness of Interventions For FatiguE in Long term conditions” (EIFFEL) (26). Study protocols have been registered on the PROSPERO database which state that the team will be conducting two systematic reviews; one to assess the effectiveness of interventions for fatigue in long term conditions (27) and the other to explore the acceptability of interventions for fatigue (28). There is potential for duplication of effort when considering acceptability of interventions, as the qualitative data may explore contextual factors and some musculoskeletal conditions are also included in their searches. However, the proposed methodology of this study is a scoping review which allows for a broader view of contextual factors across the lifespan of all chronic MSK conditions and will be gathered from multiple study designs, which is appropriate for the aims of the review to map existing knowledge. Contact has been made with the EIFFEL team to reduce risk of duplication and share knowledge.

The review has been designed in accordance with the Joanna Briggs Institute (JBI) methodology for scoping reviews (29) and will be reported using the PRISMA-ScR extension guidelines for scoping reviews (30).

### Identifying relevant studies

A preliminary search of MEDLINE in August 2023 identified the current scope and scale of the evidence base related to the scoping review. The search strategy (Appendix 1) was then developed with support from a specialist subject librarian and reviewed by stakeholders in a PPIE workshop.

The following electronic databases will be searched for research published in peer-reviewed journals from January 2007 onwards; AMED, MEDLINE, PsycINFO, CINAHLPlus (EBSCO platform), EMBASE (Ovid platform), SCOPUS and Cochrane Database. This date was chosen to correspond with the Outcome Measures in Rheumatology 8^th^ meeting (OMERACT 8) which endorsed fatigue as an addition to the ‘core set’ of outcome measures for all subsequent studies, highlighting the importance of investigating this symptom (31). There will be no restrictions on the age of participants, allowing interventions that have been used throughout the life course and highlighting any gaps in provision. A search for unpublished studies will not be conducted due to limitations of time to complete this review. The reference lists from included studies will be hand searched to check for any other relevant papers not captured in the database searching. Only evidence available in English will be reviewed and studies in other languages will be excluded due to time. All studies will be uploaded to the review software system Covidence (32).

### Selection of studies

A minimum of two independent reviewers will screen the titles and abstracts of all identified studies against the stated inclusion and exclusion criteria (Table 1). Regular meetings will be held by the team throughout the title and abstract screening process to aid understanding and reduce disagreements (33). Papers that proceed to the full text stage of screening will also be reviewed by two or more independent reviewers who will document the main reason any excluded papers do not meet the inclusion criteria. Any differences in opinion between the reviewers will be resolved by discussing the papers, with an additional independent reviewer to support mediation, as required. Any reasons for exclusion of full-text papers that do not meet the inclusion criteria will be recorded and reported in the scoping review.

**Table 1:** Inclusion and exclusion criteria.

| Inclusion criteria | Exclusion criteria |
| --- | --- |
| - Primary research study<br>- Published in a peer reviewed journal<br>- Available in English language<br>- Participants have one or more chronic MSK conditions<br>- Participants experience fatigue at baseline<br>- Published during or after 2007<br>- Describes a non-pharmacological intervention to manage MSK condition symptoms, with fatigue reduction as a primary or secondary outcome | - Reviews, protocols, opinion pieces, editorials, case series, case reports, observational cohort studies<br>- Pharmacological interventions<br>- No intervention is described<br>- Muscle fatigue rather than global fatigue is examined<br>- No data is available on factors associated with intervention success (theoretical mechanism of intervention OR characteristics of participants OR characteristics of clinicians delivering interventions) |

All primary research methodologies will be considered, including experimental and quasi-experimental study designs, before and after studies and interrupted timeseries studies, analytical observational studies, case-control studies and analytical cross-sectional studies, and descriptive observational study designs. Qualitative studies will also be considered including, for example, phenomenology, grounded theory, ethnography, qualitative description, and action research.

### Data extraction

Data from included papers will be extracted by one reviewer and checked by another using an adapted version of the JBI template for data extraction (Appendix 2). This template captures information about study participants, methods, and findings relevant to the research question (34) and has been amended to extract additional data on contextual factors including clinical characteristics, information on clinicians delivering interventions and the hypothesis behind the design of the intervention. Any disagreements that may arise between the reviewers will be resolved by consensus, with an additional reviewer to support mediation. If necessary, the authors of the included papers will be contacted for further information or data clarification.

### Data reporting and analysis

Findings will be presented in a PRISMA (Preferred Reporting Items for Systematic Reviews and Meta Analyses) flow diagram (35) to demonstrate the number of articles identified and their sources, with reasons for exclusion at full text screening summarised. All included studies will be summarised in tabular format. Further figures will be used to illustrate a map of the existing literature and any gaps highlighted by the review. Data analysis is likely to be narrative due to the broad range of study types being included. This may be further refined for use during the review process.

### Patient and public involvement statement

A patient and public involvement and engagement (PPIE) workshop was held in October 2023 during the design stage of the search strategy and review protocol. Stakeholders attending the workshop included patients living with one or more MSK conditions and clinicians from a range of professions who support patients that experience MSK related fatigue. The discussion focussed on people’s experiences of offering or being offered support to manage their fatigue and asking for comments on the proposed review. This highlighted additional intervention types and pathways that were subsequently included in the search terms (Appendix 3). It also confirmed our understanding that fatigue is a significant issue and that an overview of potential interventions and management techniques would be welcomed by patients and clinicians.

Further PPIE events are planned at key points during the project to ensure the validity of the final review. Workshops that focus on the support of adults and of children and young people will be held to discuss the initial findings following data extraction with the aim of highlighting key themes and gaps in knowledge. A further pair of workshops will be arranged following synthesis of the findings to ensure the validity of the review, discuss priorities for future research and to promote dissemination of the findings through appropriate groups. The outcome of these events and how they influence the scoping review process will be reported in the final review following the guidelines for the GRIPP2 short form reporting checklist (36), which is a tool designed to improve the reporting of public and patient involvement in research.

## Data Availability

All data produced in the present study are available upon reasonable request to the authors

## ETHICS AND DISSEMINATION

Ethical approval is not required for this scoping review. The findings of this review will be disseminated via relevant peer-reviewed journals, conference presentations and through sharing findings with relevant charities and health professionals.

## AUTHOR CONTRIBUTIONS

Project funding application by ED with support from FC, JP, GM, PM, CC, BF and ES. Drafting of protocol and search strategy by KF in discussion with ED and GY, reviewed by EE, FC, JP, GM, PM, CC, BF and ES.

## FUNDING STATEMENT

This scoping review protocol is part of the project “MusculoskelEtal faTigue acRoss the lIfe CourSe: understanding what helps and mapping what is missing (METRICS)” (reference 23140), which is jointly funded by Versus Arthritis and The Kennedy Trust.

## COMPETING INTERESTS STATEMENT

PMM has received consulting/speaker’s fees from Abbvie, BMS, Celgene, Eli Lilly, Janssen, MSD, Novartis, Orphazyme, Pfizer, Roche and UCB, all unrelated to this project. There are no competing interests in this project.

## ACKNOWLEDGEMENTS

The development of this scoping review search strategy was supported by Specialist Subject Librarian Pauline Shaw from the University of the West of England library. The review team is also grateful for the contributions of the stakeholders who attended the Patient and Public Involvement and Engagement workshop.

### Appendix 1: Search strategy overview

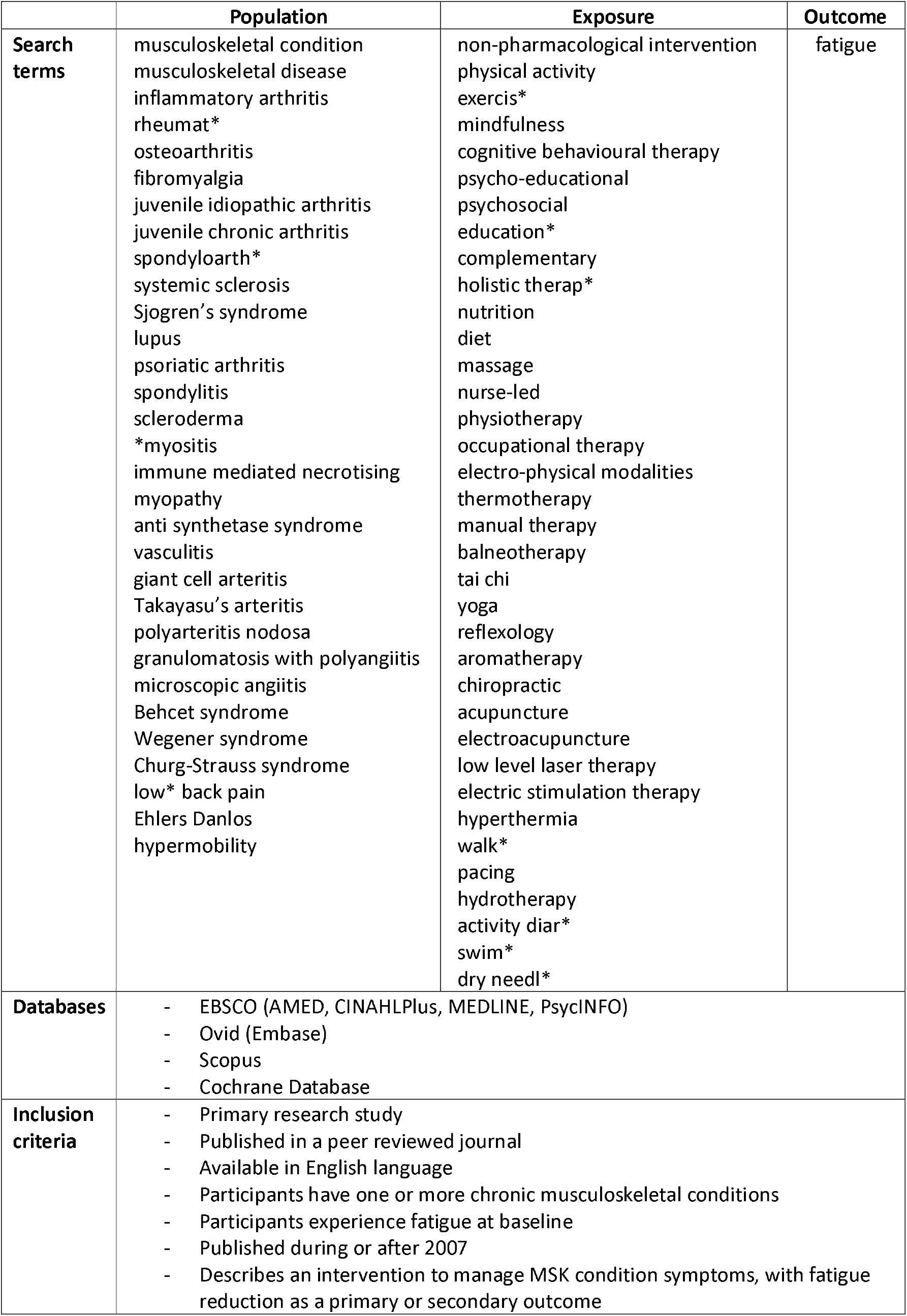
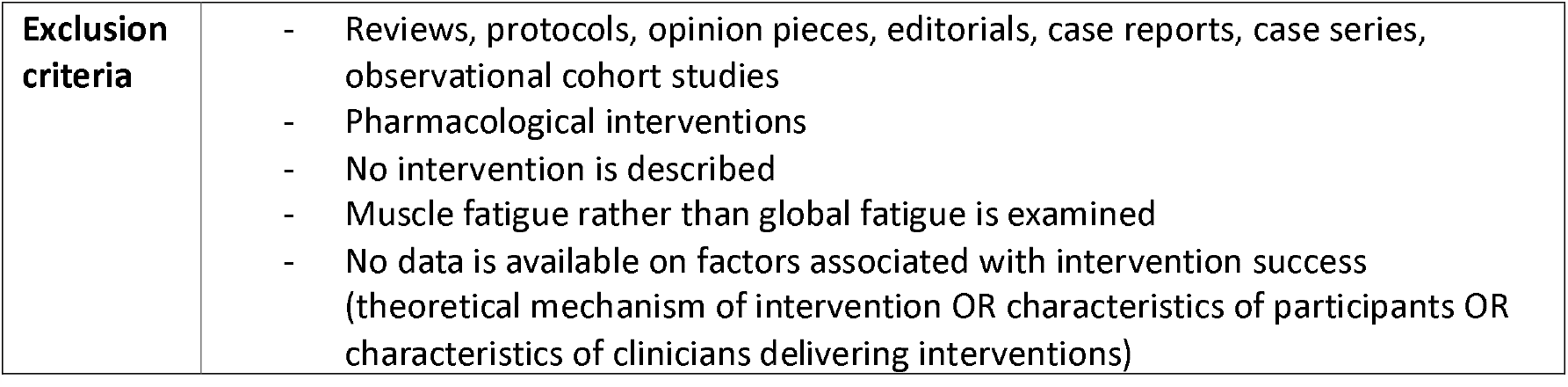

|  | Population | Exposure | Outcome |
| --- | --- | --- | --- |
| **Search terms** | musculoskeletal condition<br>musculoskeletal disease<br>inflammatory arthritis<br>rheumat*<br>osteoarthritis<br>fibromyalgia<br>juvenile idiopathic arthritis<br>juvenile chronic arthritis<br>spondyloarth*<br>systemic sclerosis<br>Sjogren's syndrome<br>lupus<br>psoriatic arthritis<br>spondylitis<br>scleroderma<br>*myositis<br>immune mediated necrotising<br>myopathy<br>anti synthetase syndrome<br>vasculitis<br>giant cell arteritis<br>Takayasu's arteritis<br>polyarteritis nodosa<br>granulomatosis with polyangiitis<br>microscopic angiitis<br>Behcet syndrome<br>Wegener syndrome<br>Churg-Strauss syndrome<br>low* back pain<br>Ehlers Danlos<br>hypermobility | non-pharmacological intervention<br>physical activity<br>exercis*<br>mindfulness<br>cognitive behavioural therapy<br>psycho-educational<br>psychosocial<br>education*<br>complementary<br>holistic therap*<br>nutrition<br>diet<br>massage<br>nurse-led<br>physiotherapy<br>occupational therapy<br>electro-physical modalities<br>thermotherapy<br>manual therapy<br>balneotherapy<br>tai chi<br>yoga<br>reflexology<br>aromatherapy<br>chiropractic<br>acupuncture<br>electroacupuncture<br>low level laser therapy<br>electric stimulation therapy<br>hyperthermia<br>walk*<br>pacing<br>hydrotherapy<br>activity diar*<br>swim*<br>dry needl* | fatigue |
| **Databases** | - EBSCO (AMED, CINAHLPlus, MEDLINE, PsycINFO)<br>- Ovid (Embase)<br>- Scopus<br>- Cochrane Database |  |  |
| **Inclusion criteria** | - Primary research study<br>- Published in a peer reviewed journal<br>- Available in English language<br>- Participants have one or more chronic musculoskeletal conditions<br>- Participants experience fatigue at baseline<br>- Published during or after 2007<br>- Describes an intervention to manage MSK condition symptoms, with fatigue reduction as a primary or secondary outcome |  |  |

|  |  |
| --- | --- |
| **Exclusion criteria** | - Reviews, protocols, opinion pieces, editorials, case reports, case series, observational cohort studies<br>- Pharmacological interventions<br>- No intervention is described<br>- Muscle fatigue rather than global fatigue is examined<br>- No data is available on factors associated with intervention success (theoretical mechanism of intervention OR characteristics of participants OR characteristics of clinicians delivering interventions) |

### Appendix 2: Amended JBI data extraction table

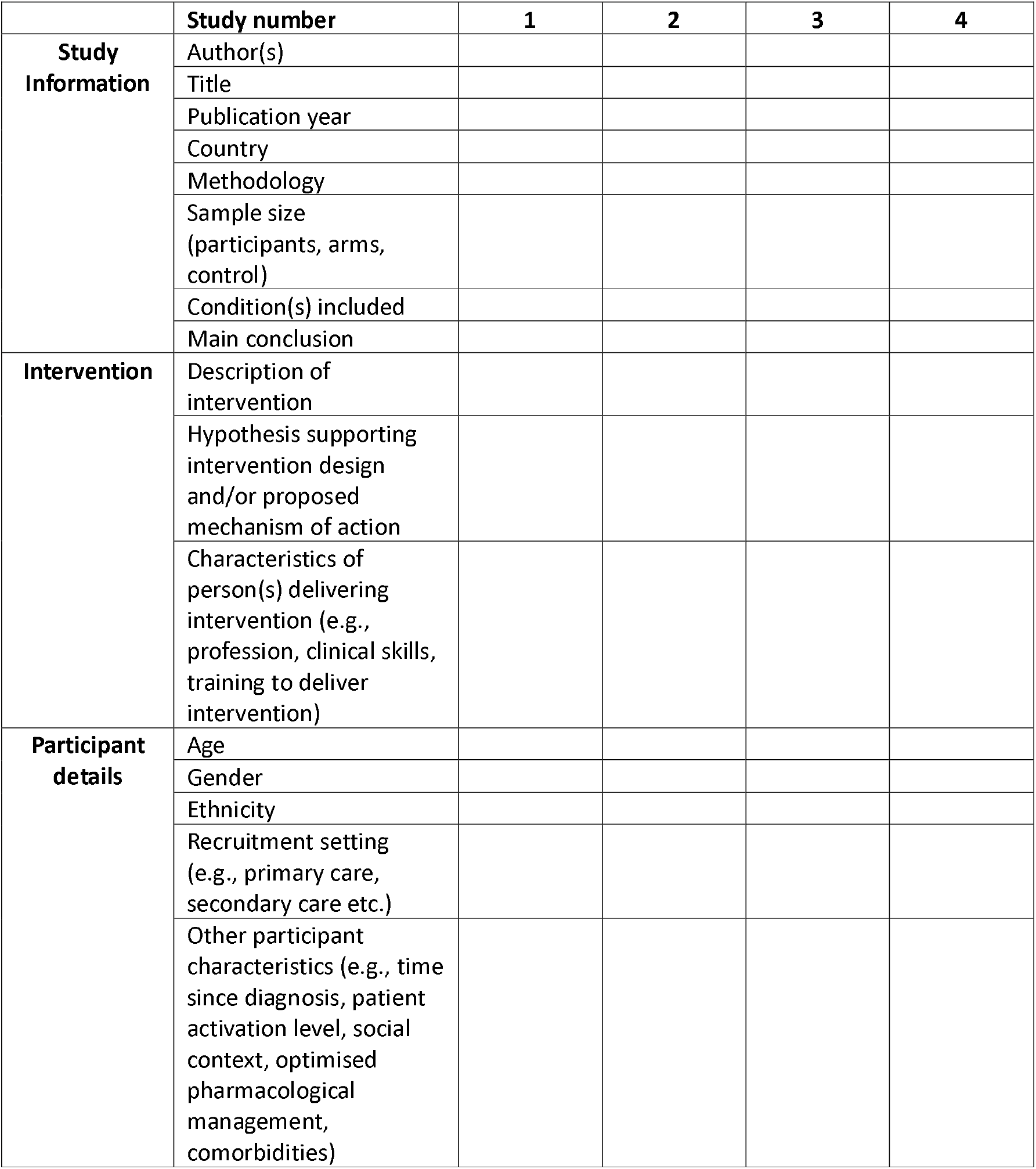

|  | Study number | 1 | 2 | 3 | 4 |
| --- | --- | --- | --- | --- | --- |
| **Study Information** | Author(s) |  |  |  |  |
|  | Title |  |  |  |  |
|  | Publication year |  |  |  |  |
|  | Country |  |  |  |  |
|  | Methodology |  |  |  |  |
|  | Sample size<br>(participants, arms,<br>control) |  |  |  |  |
|  | Condition(s) included |  |  |  |  |
|  | Main conclusion |  |  |  |  |
| **Intervention** | Description of intervention |  |  |  |  |
|  | Hypothesis supporting intervention design and/or proposed mechanism of action |  |  |  |  |
|  | Characteristics of person(s) delivering intervention (e.g., profession, clinical skills, training to deliver intervention) |  |  |  |  |
| **Participant details** | Age |  |  |  |  |
|  | Gender |  |  |  |  |
|  | Ethnicity |  |  |  |  |
|  | Recruitment setting (e.g., primary care, secondary care etc.) |  |  |  |  |
|  | Other participant characteristics (e.g., time since diagnosis, patient activation level, social context, optimised pharmacological management, comorbidities) |  |  |  |  |

### Appendix 3: Proposed search strategy

#### 3.1 EBSCO (AMED, CINAHLPlus, MEDLINE, PsycINFO)

S1 TITLE (“musculoskeletal condition” OR “musculoskeletal disease” OR “inflammatory arthritis” OR rheumat* OR osteoarthritis OR fibromyalgia OR “juvenile idiopathic arthritis” OR “juvenile chronic arthritis” OR spondyloarth* OR “systemic sclerosis” OR “Sjogren’s syndrome” OR lupus OR “psoriatic arthritis” OR spondylitis OR scleroderma OR *myositis OR “immune-mediated necrotising myopathy” OR “anti-synthetase syndrome” OR vasculitis OR “giant cell arteritis” OR “Takayasu’s arteritis” OR “polyarteritis nodosa” OR “granulomatosis with polyangiitis” OR “microscopic angiitis” OR “Behcet syndrome” OR “Wegener syndrome” OR “Churg-Strauss syndrome” OR “low* back pain” OR “Ehlers Danlos” OR hypermobility) OR ABSTRACT (“musculoskeletal condition” OR “musculoskeletal disease” OR “inflammatory arthritis” OR rheumat* OR osteoarthritis OR fibromyalgia OR “juvenile idiopathic arthritis” OR “juvenile chronic arthritis” OR spondyloarth* OR “systemic sclerosis” OR “Sjogren’s syndrome” OR lupus OR “psoriatic arthritis” OR spondylitis OR scleroderma OR *myositis OR “immune-mediated necrotising myopathy” OR “anti-synthetase syndrome” OR vasculitis OR “giant cell arteritis” OR “Takayasu’s arteritis” OR “polyarteritis nodosa” OR “granulomatosis with polyangiitis” OR “microscopic angiitis” OR “Behcet syndrome” OR “Wegener syndrome” OR “Churg-Strauss syndrome” OR “low* back pain” OR “Ehlers Danlos” OR hypermobility)

S2 TITLE (“non-pharmacological intervention” OR “physical activity” OR exercis* OR mindfulness OR “cognitive behavioural therapy” OR “psycho-educational” OR psychosocial OR education* OR complementary OR “holistic therap*” OR nutrition OR diet OR massage OR “nurse-led” OR physiotherapy OR “occupational therapy” OR “sports rehabilitation therapy” OR “electro-physical modalities” OR thermotherapy OR “manual therapy” OR balneotherapy OR “tai chi” OR yoga OR reflexology OR aromatherapy OR chiropractic OR acupuncture OR electroacupuncture OR “low level laser therapy” OR “electric stimulation therapy” OR hyperthermia OR walk* OR pacing OR hydrotherapy OR “activity diar*” OR swim* OR “dry needl*”) OR ABSTRACT (“non-pharmacological intervention” OR “physical activity” OR exercis* OR mindfulness OR “cognitive behavioural therapy” OR “psycho-educational” OR psychosocial OR education* OR complementary OR “holistic therap*” OR nutrition OR diet OR massage OR “nurse-led” OR physiotherapy OR “occupational therapy” OR “sports rehabilitation therapy” OR “electro-physical modalities” OR thermotherapy OR “manual therapy” OR balneotherapy OR “tai chi” OR yoga OR reflexology OR aromatherapy OR chiropractic OR acupuncture OR electroacupuncture OR “low level laser therapy” OR “electric stimulation therapy” OR hyperthermia OR walk* OR pacing OR hydrotherapy OR “activity diar*” OR swim* OR “dry needl*”)

S3 TITLE (fatigue) OR ABSTRACT (fatigue)

S4 S1 AND S2 AND S3

S5 Limit S5 to;

- Publication date during or after 2007
- English language
- Peer reviewed

#### 3.2 Ovid (EMBASE)

#1 (musculoskeletal condition OR musculoskeletal disease OR inflammatory arthritis OR rheumat* OR osteoarthritis OR fibromyalgia OR juvenile idiopathic arthritis OR spondyloarth* OR systemic sclerosis OR Sjogrens syndrome OR lupus OR psoriatic arthritis OR spondylitis OR scleroderma OR myositis OR immune-mediated necrotising myopathy OR anti-synthetase syndrome OR vasculitis OR giant cell arteritis OR Takayasus arteritis OR polyarteritis nodosa OR granulomatosis with polyangiitis OR microscopic angiitis OR Behcet syndrome OR Wegener syndrome OR Churg-Strauss syndrome OR low* back pain OR Ehlers Danlos OR hypermobility).ti OR (musculoskeletal condition OR musculoskeletal disease OR inflammatory arthritis OR rheumat* OR osteoarthritis OR fibromyalgia OR juvenile idiopathic arthritis OR spondyloarth* OR systemic sclerosis OR Sjogrens syndrome OR lupus OR psoriatic arthritis OR spondylitis OR scleroderma OR myositis OR immune-mediated necrotising myopathy OR anti-synthetase syndrome OR vasculitis OR giant cell arteritis OR Takayasus arteritis OR polyarteritis nodosa OR granulomatosis with polyangiitis OR microscopic angiitis OR Behcet syndrome OR Wegener syndrome OR Churg-Strauss syndrome OR low* back pain OR Ehlers Danlos OR hypermobility).ab.

#2 Limit 1 to (English language and yr=“2007”)

#3 (fatigue).ti. or (fatigue).ab.

#4 Limit 2 to (English language and yr=“2007”)

#5 (non-pharmacological intervention OR physical activity or exercis* OR mindfulness OR cognitive behavioural therapy OR psycho-educational OR psychosocial OR education* OR complementary OR holistic therp* OR nutrition OR diet OR massage OR nurse-led OR physiotherapy OR occupational therapy OR sports rehabilitation therapy OR electro-physical modalities OR thermotherapy OR manual therapy OR balneotherapy OR tai chi OR yoga OR reflexology OR aromatherapy OR chiropractic OR acupuncture OR electroacupuncture OR low level laser therapy OR electric stimulation therapy OR hyperthermia OR walk* OR pacing OR hydrotherapy OR activity diar* OR swim* OR dry needl*).ti. OR (non-pharmacological intervention OR physical activity or exercis* OR mindfulness OR cognitive behavioural therapy OR psycho-educational OR psychosocial OR education* OR complementary OR holistic therp* OR nutrition OR diet OR massage OR nurse-led OR physiotherapy OR occupational therapy OR sports rehabilitation therapy OR electro-physical modalities OR thermotherapy OR manual therapy OR balneotherapy OR tai chi OR yoga OR reflexology OR aromatherapy OR chiropractic OR acupuncture OR electroacupuncture OR low level laser therapy OR electric stimulation therapy OR hyperthermia OR walk* OR pacing OR hydrotherapy OR activity diar* OR swim* OR dry needl*).ab.

#6 Limit 5 to (English language and yr=“2007”)

#7 2 and 4 and 6

#### 3.3 SCOPUS

#1 (TITLE-ABS(“non pharmacological intervention” OR “physical activity” OR exercis* OR mindfulness OR “cognitive behavioural therapy” OR psycho-educational OR psychosocial OR education* OR complementary OR “holistic therap*” OR nutrition OR diet OR massage OR “nurse-led” OR physiotherapy OR “occupational therapy” OR “sports rehabilitation therapy” OR “electro-physical modalities” OR thermotherapy OR “manual therapy” OR balneotherapy OR “tai chi” OR yoga OR reflexology OR aromatherapy OR chiropractic OR acupuncture OR electroacupuncture OR “low level laser therapy” OR “electric stimulation therapy” OR hyperthermia OR walk* OR pacing OR hydrotherapy OR “activity diar*” OR swim* OR “dry needl*”)

#2 Limit #1 to:

- Publication during or after 2007
- English language

#3 TITLE-ABS(fatigue)

#4 Limit #3 to:

- Publication during or after 2007
- English language

#5 TITLE-ABS(“musculoskeletal condition” OR “musculoskeletal disease” OR “inflammatory arthritis” OR rheumat* OR osteoarthritis OR fibromyalgia OR “juvenile idiopathic arthritis” OR spondyloarth* OR “systemic sclerosis” OR “Sjogrens syndrome” OR lupus OR “psoriatic arthritis” OR spondylitis OR scleroderma OR *myositis OR “immune-mediated necrotising myopathy” OR “anti-synthetase syndrome” OR vasculitis OR “giant cell arteritis” OR “Takayasus arteritis” OR “polyarteritis nodosa” OR “granulomatosis polyangiitis” OR “microscopic angiitis” OR “Behcet syndrome” OR “Wegener syndrome” OR “Churg-Strauss syndrome” OR “low* back pain” OR “Ehlers Danlos” OR hypermobility)

#6 Limit #5 to:

- Publication during or after 2007
- English language

#7 #2 AND #4 AND #6

#### 3.4 COCHRANE LIBRARY DATABASE

ID Search terms Results

#1 (“non pharmacological intervention” OR “physical activity” OR exercis* OR mindfulness OR “cognitive behavioural therapy” OR psycho-educational OR psychosocial OR education* OR complementary OR “holistic therap*” OR nutrition OR diet OR massage OR “nurse-led” OR physiotherapy OR “occupational therapy” OR “sports rehabilitation therapy” OR “electro-physical modalities” OR thermotherapy OR “manual therapy” OR balneotherapy OR “tai chi” OR yoga OR reflexology OR aromatherapy OR chiropractic OR acupuncture OR electroacupuncture OR “low level laser therapy” OR “electric stimulation therapy” OR hyperthermia OR walk* OR pacing OR hydrotherapy OR “activity diar*” OR swim* OR “dry needl*”):ti,ab

#2 (fatigue):ti,ab

#3 (“musculoskeletal condition” OR “musculoskeletal disease” OR “inflammatory arthritis” OR rheumat* OR osteoarthritis OR fibromyalgia OR “juvenile idiopathic arthritis” OR spondyloarth* OR “systemic sclerosis” OR “Sjogren’s syndrome” OR lupus OR “psoriatic arthritis” OR spondylitis OR scleroderma OR *myositis OR “immune-mediated necrotising myopathy” OR “anti-synthetase syndrome” OR vasculitis OR “giant cell arteritis” OR “Takayasu’s arteritis” OR “polyarteritis nodosa” OR “granulomatosis with polyangiitis” OR “microscopic angiitis” OR “Behcet syndrome” OR “Wegener syndrome” OR “Churg-Strauss syndrome” OR “low* back pain” OR “Ehlers Danlos” OR hypermobility):ti,ab

#4 #1 AND #2 AND #3

#5 Limit to:

- Publication date from Jan 2007 to Oct 2023
- English language

## Notes

### Competing Interest Statement

PMM has received consulting/speaker fees from Abbvie, BMS, Celgene, Eli Lilly, Janssen, MSD, Novartis, Orphazyme, Pfizer, Roche and UCB, all unrelated to this project. There are no competing interests in this project.

### Clinical Protocols

https://osf.io/s7c6a/

